# SARS-CoV-2 Omicron symptomatic infections in previously infected or vaccinated South African healthcare workers

**DOI:** 10.1101/2022.02.04.22270480

**Authors:** Marta C. Nunes, Sthembile Sibanda, Vicky L. Baillie, Gaurav Kwatra, Ricardo Aguas, Shabir A. Madhi, the Wits VIDA HCW Study Group

## Abstract

We investigated Omicron infections among healthcare workers (HCW) presenting with symptoms of SARS-CoV-2 infection and evaluated the protective effect of vaccination or prior infection.

Between 24^th^ November and 31^st^ December 2021, HCW in Johannesburg, South Africa, were tested for SARS-CoV-2 infection by Nucleic Acid Amplification Test (NAAT). Blood samples collected either at the symptomatic visit or within 3-months prior, were tested for spike protein immunoglobulin G (IgG).

Overall, 433 symptomatic HCW were included in the analysis, with 190 (43.9%) having an Omicron infection; 69 (16.7%) were unvaccinated and 270 (62.4%) received a single dose of Ad26.COV.2 vaccine. There was no difference in the odds of identifying Omicron between unvaccinated and Ad26.COV.2 vaccinated HCW (adjusted odds ratio [aOR] 0.81, 95% confidence interval [CI]: 0.46, 1.43). One-hundred and fifty-four (35.3%) HCW had at least one SARS-CoV-2 NAAT-confirmed prior infection; these had lower odds of Omicron infection compared with those without past infection (aOR 0.55, 95%CI: 0.36, 0.84). Anti-spike IgG concentration of 1549 binding antibody unit/mL was suggestive of significant reduction in the risk of symptomatic Omicron infection.

We found high reinfection and vaccine breakthrough infection rates with the Omicron variant among HCW. Prior infection and high anti-spike IgG concentration were protective against Omicron infection.

## Introduction

The Omicron (B.1.1.529/21K) severe acute respiratory syndrome coronavirus-2 (SARS-CoV-2) variant was reported in South Africa on 25^th^ November 2021, following investigation of a rapid increase in coronavirus disease (COVID-19) cases in the Gauteng province, and identification of a spike gene target failure (SGTF) on the Taqpath assay (ThermoFisher™), which also includes two other gene targets. In December 2021, the Omicron variant constituted 98% of all SARS-CoV-2 infections in South Africa, and has now spread globally [1].

Healthcare workers (HCW) in South Africa were offered the Ad26.COV.2 COVID-19 vaccine as part of the Sisonke trial from 17^th^ February 2021 as a single dose schedule, and subsequently a booster dose was offered since 8^th^ November 2021 [2]. From May 2021, HCW could also access the BNT162b2 COVID-19 vaccine as part of the national vaccine rollout in South Africa.

Here we describe the Omicron infections among HCW who presented with symptoms suggestive of SARS-CoV-2 infection from 24^th^ November to 31^st^ December 2021. We also detail breakthrough infections in vaccinated HCW and reinfections in previous Nucleic Acid Amplification Test (NAAT)-confirmed SARS-CoV-2 cases. In addition, blood samples collected either at the symptomatic visit or in the 3-months prior, were tested for full-length SARS-CoV-2 spike protein immunoglobulin G (IgG) to assess the potential protective effect of these antibodies.

## Methods

### Study design

Healthcare workers (HCW) working at Chris Hani Baragwanath Academic Hospital (CHBAH), in Johannesburg, Gauteng province, South Africa were enrolled from April to July 2020, into a longitudinal cohort surveillance study, this cohort has been previously described [3]. Due to participants discontinuing the study, enrolments into the longitudinal cohort were re-started on 16^th^ February 2021 and closed on 10^th^ August 2021. Among the longitudinal cohort participants, routine study visits (every 1 to 2 weeks for nasal swab collection and approximately every 4 weeks for nasal swab and venous blood collection) and visits when COVID-like symptoms are present are still ongoing.

From 22^nd^ June 2021 HCW from CHBAH, not enrolled into the longitudinal cohort, who presented with COVID-like symptoms could enrol into a test negative case-control (TNC) study and be tested for SARS-CoV-2 infection by Nucleic Acid Amplification Test (NAAT) on nasal swab. On 14^th^ December 2021 this was expanded to two other Johannesburg hospitals: Charlotte Maxeke Johannesburg Academic Hospital (CMJAH) and Helen Joseph Hospital (HJH). HCW enrolled into the TNC could present for multiple study visits. Enrolments are still ongoing at the three hospitals. HCW in the longitudinal cohort who were investigated for symptomatic illness were also eligible for inclusion in the TNC study.

Symptoms considered to be consistent with COVID-19 included any of the following: fever/feeling feverish, cough, sore throat, rhinitis, myalgia, shortness of breath, acute gastroenteritis/vomiting/nausea, impaired sense of smell or taste, fatigue or headache. If a HCW had multiple symptomatic study visits between 24^th^ November and 31^st^ December 2021 only the visit with a NAAT SARS-CoV-2 positive result was included, or the first symptomatic visit if no NAAT positive result.

Demographic, health and behavioural questionnaires, collected personal information including COVID-19 vaccination history; previous SARS-CoV-2 infection was determined by documented NAAT positivity in the cohort participants or self-reporting.

### Laboratory methods

Total nucleic acids were extracted from nasal swabs using an automated NucliSENS-easyMAG nucleic acid extraction platform. NAAT was performed using the TaqPath COVID-19 diagnostic test from ThermoFisher that uses a triple-target (orf1ab, N gene, spike gene) design. Results were classified as positive for SARS-CoV-2 when the 3 targets or when both orf1ab and N gene had cycle threshold (Ct) values <37 and inconclusive if only one target was detected with Ct values <37. Results were classified as Omicron variant when orf1ab and N gene were detected but not the spike gene (SGTF).

Serum or plasma samples were collected at approximately monthly intervals from the longitudinal cohort participants. Participants enrolled into the TNC study had blood samples collected at enrolment. SARS-CoV-2 full-length spike protein immunoglobulin G (IgG) was measured by a quantitative assay on the Luminex platform as described [4, 5]. The assay was evaluated for detection of antibodies against SARS-CoV-2 using COVID-19 convalescent plasma panel NIBSC 20/118. Based on differences in IgG titers from pre-COVID-19 and baseline samples when compared to post-infection samples of participants who were SARS-CoV-2 NAAT positive, 32 binding antibody unit (BAU)/mL was selected as the threshold value indicative of seropositivity for full-length spike.

### Statistical analysis

Participants’ characteristics were described as percentages or means with standard deviations (SD) or median with interquartile range (IQR) and compared between NAAT-confirmed Omicron infected and uninfected HCW. The association between Omicron infection, and vaccination or previous SARS-CoV-2 infection, was estimated by univariate and multivariate logistic regression. Participants were considered fully-vaccinated if they received at least one dose of Ad26.COV.2 vaccine ≥ 14 days before the symptomatic visit or two doses of BNT162b2 vaccine, with the last dose ≥ 14 days before the symptomatic visit. Prior SARS-CoV-2 infections were categorized under the three previous pandemic waves: April to October 2020, November 2020 to April 2021 and May to September 2021. Differences in geometric mean units for spike IgG between NAAT-confirmed Omicron infected and uninfected HCW were analysed on log_10_-transformed data.

A recursive partitioning approach was performed in the form of conditional inference tree. This method is particularly good at finding conditional thresholds in covariates by means of significance tests. Significance tests of the null hypothesis, that Omicron infection risk is identical on either side of the proposed threshold, are performed at each node of the tree in a recursive way. The final tree outlines all the splits for which the null hypothesis was rejected, i.e., for which a difference in outcome is statistically significant with 90% confidence across merging branches. With this type of conditional partitioning, a training set can be used to find the relevant splits, after which a validation portion of the data can be used to evaluate the method’s predictive power [6]. This analysis was performed using the rpart, party, and caret R packages.

For the purposes of the analyses presented throughout, serological measurements were restricted to blood samples collected within 3 months of symptomatic visits. If multiple samples were available, the sample collected closest to the symptomatic visit was used. Participants were excluded from the serology analysis if they received any vaccine between the last blood draw and the symptomatic visit or if the last blood draw was <14 days after the last vaccination. Since IgG antibodies after natural infection among vaccinated and previous infected individuals can rise quickly, we did a sub-analysis excluding the samples collected on the day of the symptomatic visit.

## Results

The first case of SARS-CoV-2 with SGTF among HCW in our study was detected on 24^th^ November 2021, prior to which the last confirmed SARS-CoV-2 infection was on 20^th^ September 2021. From 24^th^ November to 31^st^ December 2021, 445 HCW had at least one symptomatic visit where nasal swabs were collected and tested by Taqpath NAAT for SARS-CoV-2 infection. Nine HCW had inconclusive results and were excluded from the analysis, and all but three (also excluded from the analysis) of the SARS-CoV-2 infections detected during this period had NAAT results with SGTF and putatively were Omicron variant.

Among the 433 symptomatic HCW included in the analysis, 190 (43.9%) had a positive NAAT. Overall, 270 (62.4%) received a single dose of Ad26.COV.2 vaccine (median 280-days; interquartile range [IQR]: 257, 287), and 49 (11.8%) received a booster dose (median of 22-days; IQR: 18, 33) ≥ 14-days before the symptomatic visit. Only 26 (6.3%) HCW received two doses of BNT162b2 ≥ 14-days before the visit, and 69 (16.7%) were unvaccinated. Vaccination coverage was similar among HCW with symptomatic illness in whom Omicron was and was not identified (Table 1). Also, there was no difference in the odds of identifying Omicron between unvaccinated and vaccinated HCW, although the numbers for BNT162b recipients were low (Table 2).

**Table 1.**
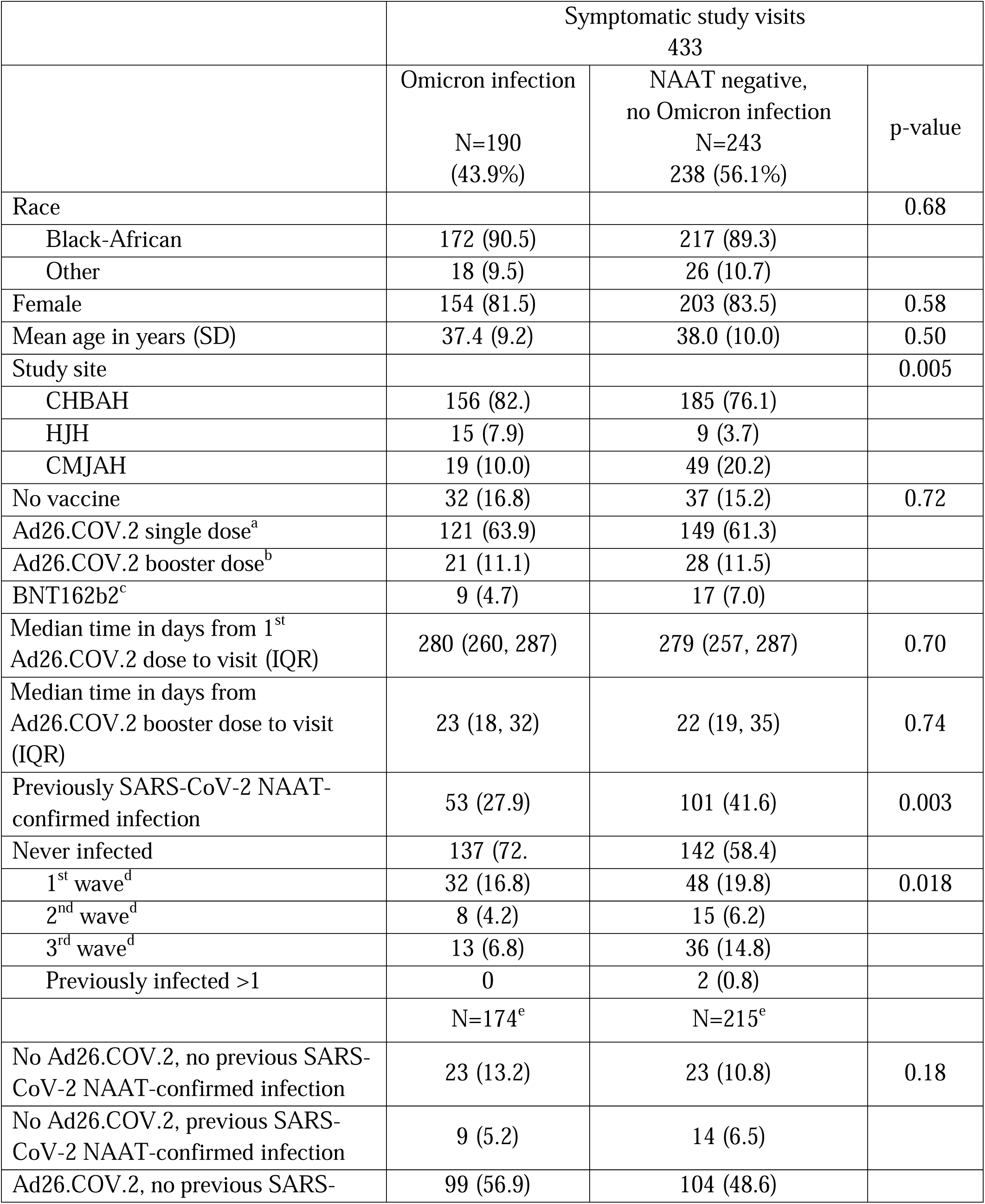
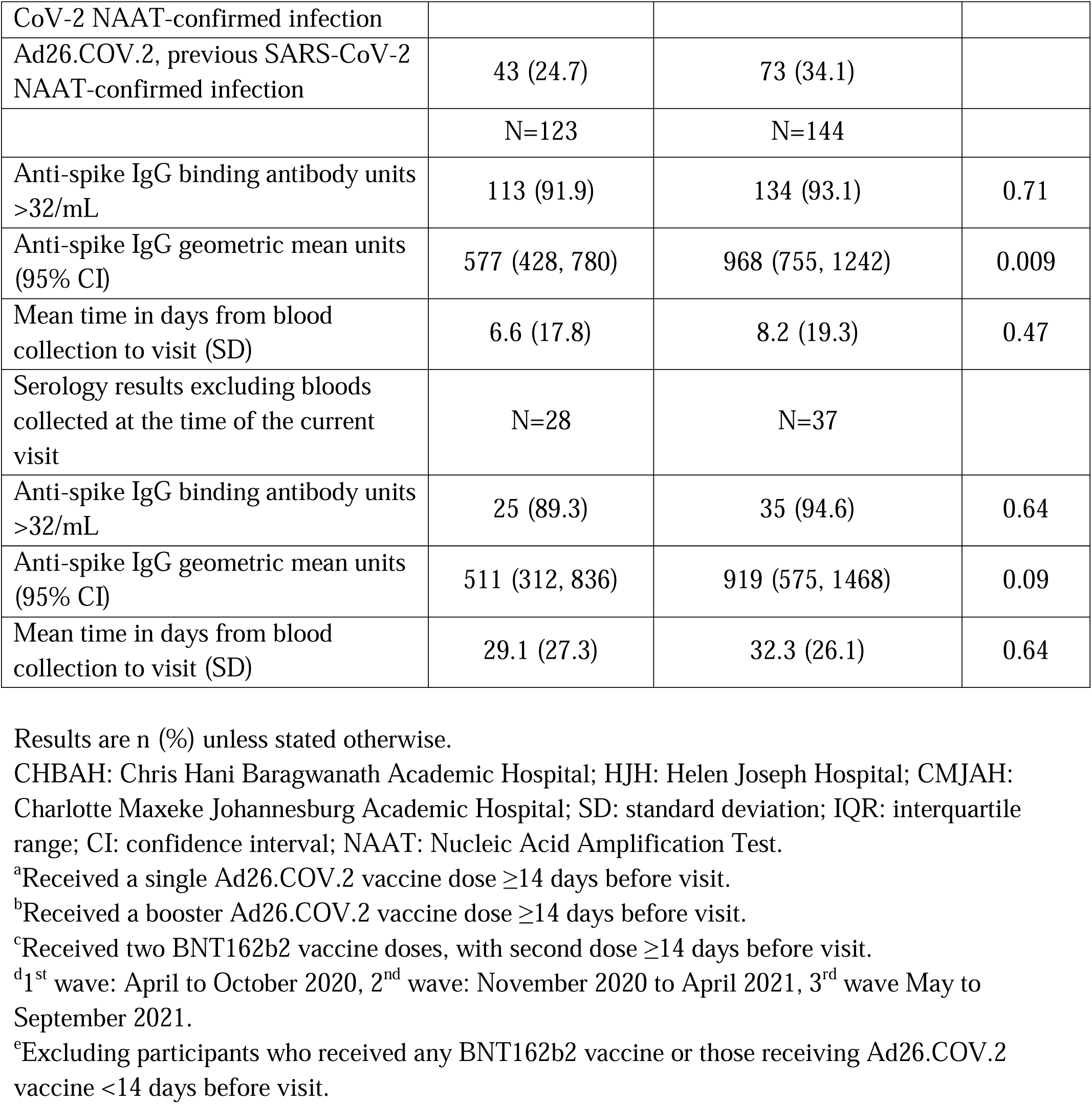
Healthcare workers presenting for at least one symptomatic study visit between 24^th^ November and 31^st^ December 2021

**Table 2.**
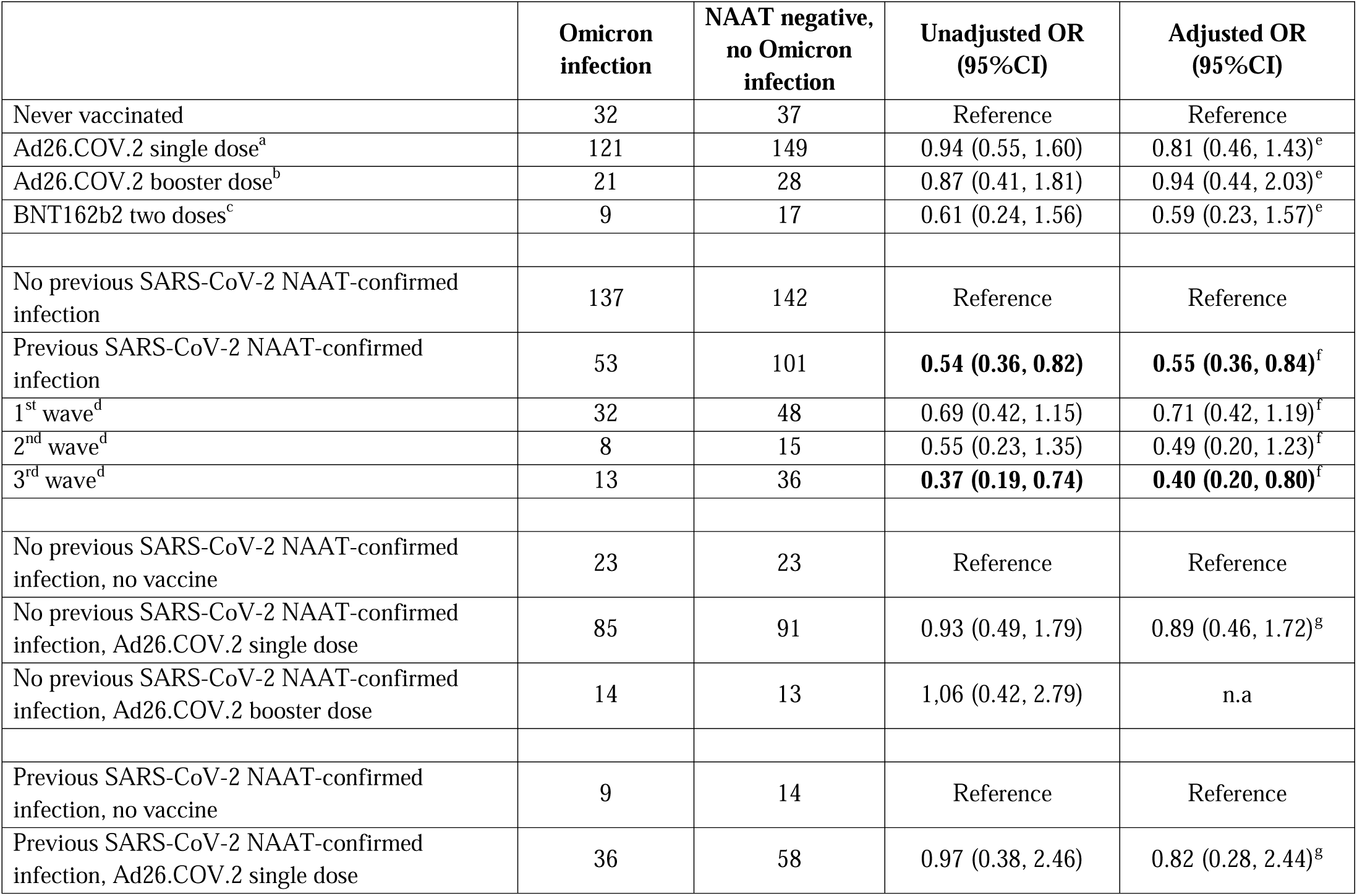
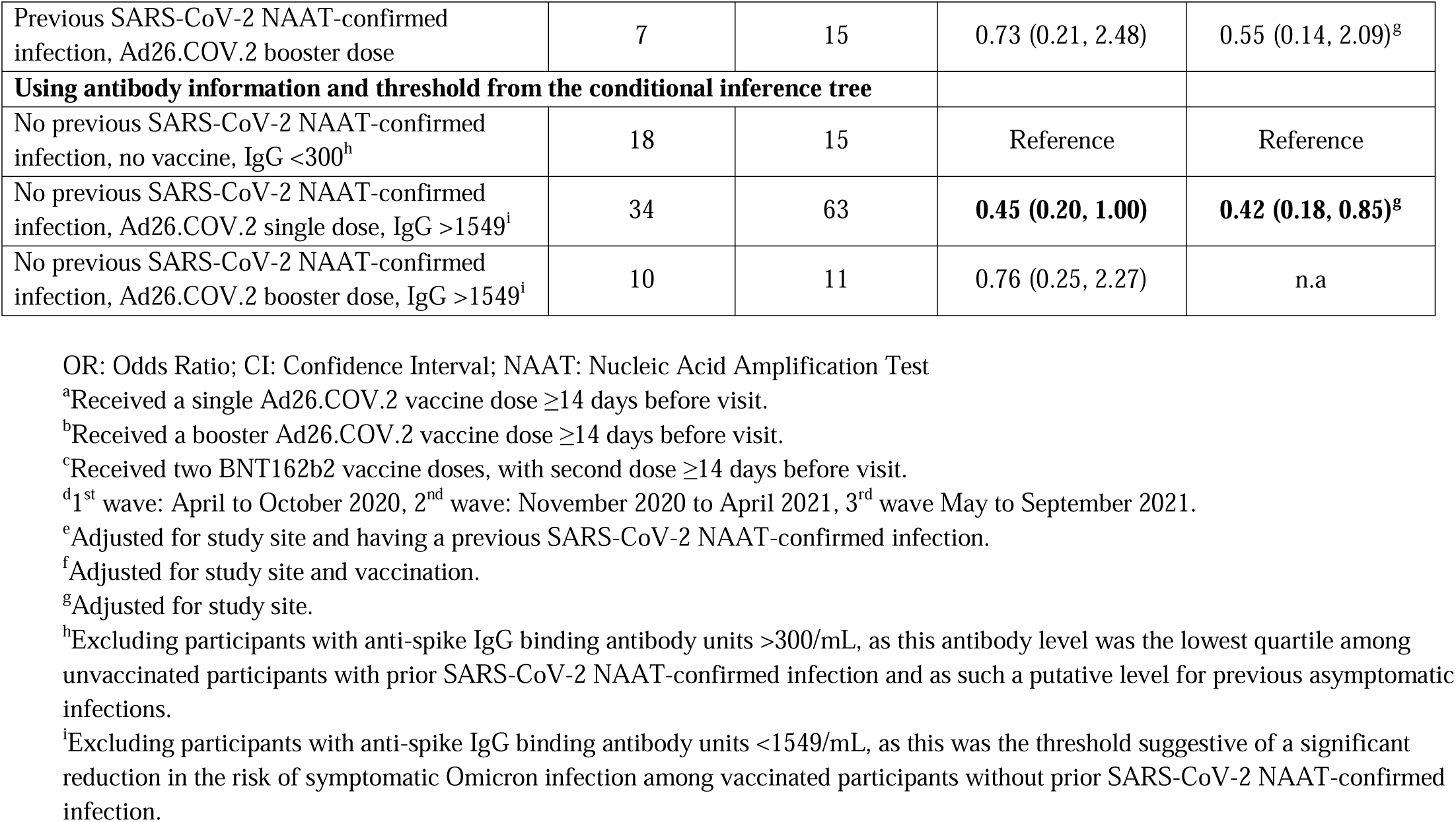
Protection against Omicron infection by vaccination or previous SARS-CoV-2 NAAT-confirmed infection

|  | **Omicron infection** | **NAAT negative, no Omicron infection** | **Unadjusted OR (95%CI)** | **Adjusted OR (95%CI)** |
| --- | --- | --- | --- | --- |
| Never vaccinated | 32 | 37 | Reference | Reference |
| Ad26.COV.2 single dose<sup>a</sup> | 121 | 149 | 0.94 (0.55, 1.60) | 0.81 (0.46, 1.43)<sup>e</sup> |
| Ad26.COV.2 booster dose<sup>b</sup> | 21 | 28 | 0.87 (0.41, 1.81) | 0.94 (0.44, 2.03)<sup>e</sup> |
| BNT162b2 two doses<sup>c</sup> | 9 | 17 | 0.61 (0.24, 1.56) | 0.59 (0.23, 1.57)<sup>e</sup> |
| No previous SARS-CoV-2 NAAT-confirmed infection | 137 | 142 | Reference | Reference |
| Previous SARS-CoV-2 NAAT-confirmed infection | 53 | 101 | **0.54 (0.36, 0.82)** | **0.55 (0.36, 0.84)<sup>f</sup>** |
| 1<sup>st</sup> wave<sup>d</sup> | 32 | 48 | 0.69 (0.42, 1.15) | 0.71 (0.42, 1.19)<sup>f</sup> |
| 2<sup>nd</sup> wave<sup>d</sup> | 8 | 15 | 0.55 (0.23, 1.35) | 0.49 (0.20, 1.23)<sup>f</sup> |
| 3<sup>rd</sup> wave<sup>d</sup> | 13 | 36 | **0.37 (0.19, 0.74)** | **0.40 (0.20, 0.80)<sup>f</sup>** |
| No previous SARS-CoV-2 NAAT-confirmed infection, no vaccine | 23 | 23 | Reference | Reference |
| No previous SARS-CoV-2 NAAT-confirmed infection, Ad26.COV.2 single dose | 85 | 91 | 0.93 (0.49, 1.79) | 0.89 (0.46, 1.72)<sup>g</sup> |
| No previous SARS-CoV-2 NAAT-confirmed infection, Ad26.COV.2 booster dose | 14 | 13 | 1.06 (0.42, 2.79) | n.a |
| Previous SARS-CoV-2 NAAT-confirmed infection, no vaccine | 9 | 14 | Reference | Reference |
| Previous SARS-CoV-2 NAAT-confirmed infection, Ad26.COV.2 single dose | 36 | 58 | 0.97 (0.38, 2.46) | 0.82 (0.28, 2.44)<sup>g</sup> |
| Previous SARS-CoV-2 NAAT-confirmed infection, Ad26.COV.2 booster dose | 7 | 15 | 0.73 (0.21, 2.48) | 0.55 (0.14, 2.09)<sup>g</sup> |
| **Using antibody information and threshold from the conditional inference tree** |  |  |  |  |
| No previous SARS-CoV-2 NAAT-confirmed infection, no vaccine, IgG <300<sup>h</sup> | 18 | 15 | Reference | Reference |
| No previous SARS-CoV-2 NAAT-confirmed infection, Ad26.COV.2 single dose, IgG >1549<sup>i</sup> | 34 | 63 | **0.45 (0.20, 1.00)** | **0.42 (0.18, 0.85)<sup>g</sup>** |
| No previous SARS-CoV-2 NAAT-confirmed infection, Ad26.COV.2 booster dose, IgG >1549<sup>i</sup> | 10 | 11 | 0.76 (0.25, 2.27) | n.a |

OR: Odds Ratio; CI: Confidence Interval; NAAT: Nucleic Acid Amplification Test
<sup>a</sup>Received a single Ad26.COV.2 vaccine dose $\geq 14$ days before visit.
<sup>b</sup>Received a booster Ad26.COV.2 vaccine dose $\geq 14$ days before visit.
<sup>c</sup>Received two BNT162b2 vaccine doses, with second dose $\geq 14$ days before visit.
<sup>d</sup>1<sup>st</sup> wave: April to October 2020, 2<sup>nd</sup> wave: November 2020 to April 2021, 3<sup>rd</sup> wave May to September 2021.
<sup>e</sup>Adjusted for study site and having a previous SARS-CoV-2 NAAT-confirmed infection.
<sup>f</sup>Adjusted for study site and vaccination.
<sup>g</sup>Adjusted for study site.
<sup>h</sup>Excluding participants with anti-spike IgG binding antibody units >300/mL, as this antibody level was the lowest quartile among unvaccinated participants with prior SARS-CoV-2 NAAT-confirmed infection and as such a putative level for previous asymptomatic infections.

Overall, 154 (35.6%) HCW had at least one SARS-CoV-2 NAAT-confirmed infection prior to November 2021, 53 (34.4%) of whom were reinfected with Omicron; compared with 137 Omicron infections among the 279 (49.1% p=0.003) HCW without previous NAAT-confirmed infection. Participants with previous NAAT-confirmed infection had lower odds of Omicron infection compared with those without past infection (adjusted odds ratio [aOR] 0.55, 95% confidence interval [CI]: 0.36, 0.84). Stratifying by timing of previous infection, infection during the preceding third wave was associated with lower odds of symptomatic Omicron illness relative to HCW without any previous NAAT-confirmed infection (aOR 0.40, 95%CI: 0.20, 0.80); likewise, individuals who were infected during the second wave had similar lower odds of being infected with Omicron during the study period (aOR 0.49, 95%CI: 0.20, 1.23), although not significant (Table 2).

Anti-spike IgG geometric mean units (measured in 267 participants) were lower in HCW who eventually had an Omicron infection compared with those who never tested positive (577 binding antibody unit [BAU]/mL, vs. 968BAU/mL, p=0.009) (Table 1). Excluding blood samples collected at the time of the current visit, a similar trend in IgG levels was observed (Table 1).

To further investigate which combinations of covariates significantly modulate Omicron infection, a conditional inference tree was built (Figure-A). Significance was detected in previous SARS-CoV-2 NAAT-confirmed cases and those with spike IgG levels >1549BAU/mL (Figure-B), each with only 33% probability of infection. The boxplots in Figure-C represent the anti-spike IgG levels by prior SARS-CoV-2 NAAT-confirmed infection and vaccination status. Overall, IgG concentrations were higher among HCW with prior infection (p=0.00015), and in the group not previously infected in those with more vaccine doses (p=0.000057). A lower significance was detected among the groups with different vaccination status for those who had a prior confirmed SARS-CoV-2 infection (p=0.038).

**Figure.**
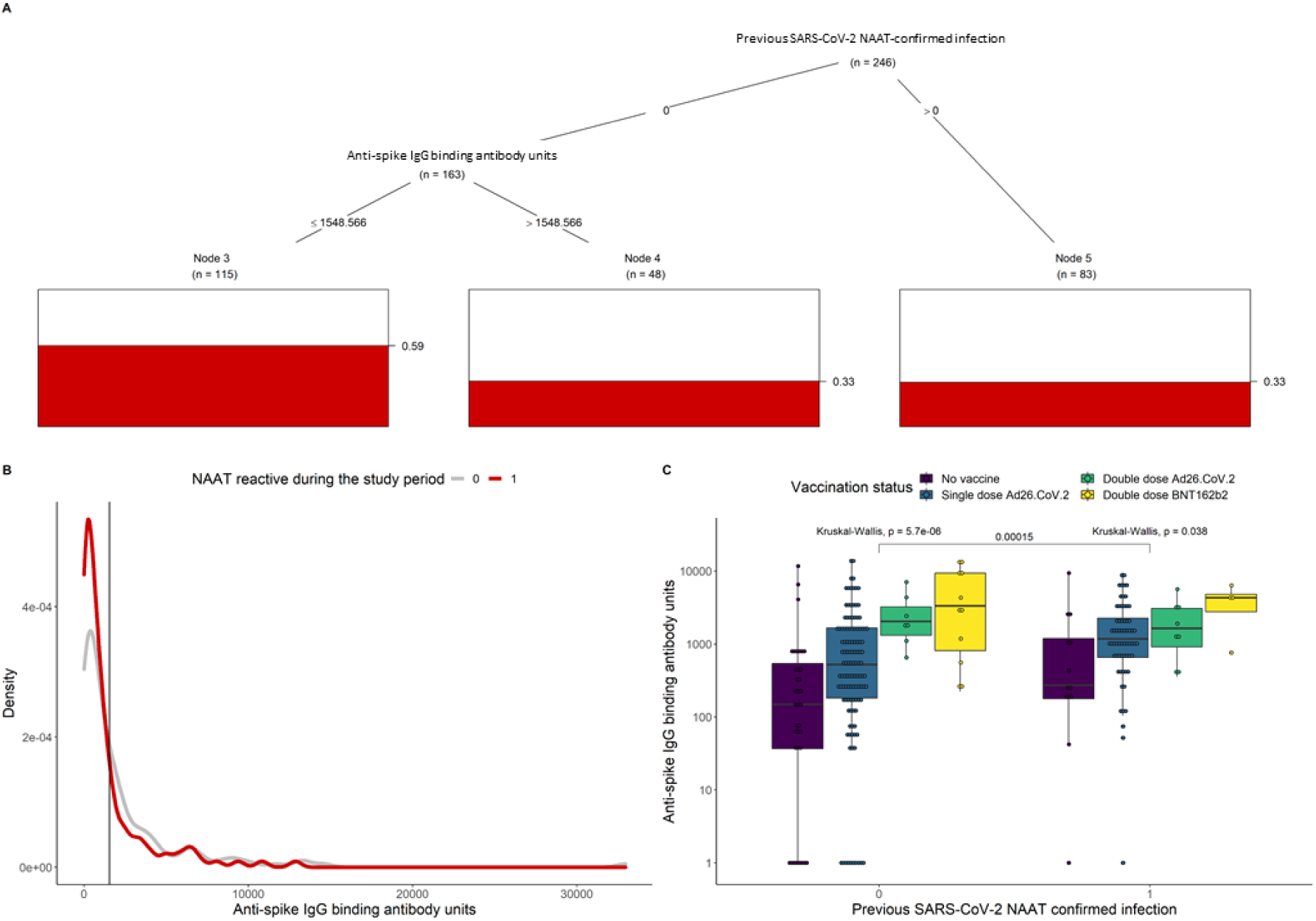
Conditional inference of Omicron infection probability and anti-spike IgG levels by prior SARS-CoV-2 NAAT-confirmed infection (A) Inferred significant splits in previous SARS-CoV-2 NAAT-confirmed cases and spike IgG levels impact on the probability of having an Omicron infection during the study period (indicated by the red bars). The tree was generated from a training set comprised of 90% of all visits with a known serological result. The algorithms infection predictive power was measured to be 72% in the remaining 10% of the data. (B) Antibody density distributions for participants with either NAAT-confirmed Omicron infection (red line) or no infection (grey line) during the study period. The vertical black line corresponds to the threshold 1549 anti-spike IgG binding units that emerged from the analysis on panel A. (C) Anti-spike IgG levels by prior SARS-CoV-2 NAAT-confirmed infection and vaccination status. Kruskal-Wallis tests indicate significant differences in IgG concentrations between participants with different levels of prior infection (p=0.00015), and with more doses of vaccination in those who were not previously exposed (p=0.000057). A lower significance in differences in IgG concentration for groups with different vaccination status for those who had a prior confirmed SARS-CoV-2 infection (p=0.038).

## Discussion

We show that prior SARS-CoV-2 infection prevented symptomatic reinfection with Omicron at 45%-60%, what is consistent with results from a large population study in Qatar [7]. Although it has been suggested that Omicron is evasive to neutralizing antibodies induced by natural infection from previous variants or vaccine-elicited[8], we show that HCW who were not infected by Omicron during the five week period of this analysis had higher concentration of anti-spike IgG prior the symptomatic visit, with >1549BAU/mL being the threshold suggestive of significant reduction in the risk of symptomatic Omicron infection. This might be due to some residual neutralization activity, or that anti-spike IgG recognizes the virus beyond neutralization via Fc-effector mechanisms as recently suggested [9]. Protection can be achieved by prior infection (irrespective of the vaccination status, Table) or by recent vaccination, with participants who were unvaccinated or those who received a single dose of Ad26.COV.2, most of whom more than eight months prior, showing significantly lower levels of anti-spike IgG.

A limitation of our study was that prior infection was assessed by NAAT only, with some unvaccinated participants with no previous NAAT-confirmed infection being seropositive for anti-spike antibody, demonstrating exposure to SARS-CoV-2 before blood collection. A high SARS-CoV-2 seropositivity in South Africa prior to the Omicron wave, has actually been suggested as a plausible explanation for the disconnection between hospitalization/death rates and infection rates associated with Omicron in the country [4].

We found high reinfection and vaccine breakthrough infection rates with the Omicron variant among HCW at three hospitals in Johannesburg, South Africa. Inherently, either natural infection or vaccination elicits immune responses that decays over time, with the specificity (cross-immunity), quality (neutralization) and magnitude (absolute amount) of circulating antibodies determining the likelihood of future symptomatic infections. Although a study from the United Kingdom showed limited protection against symptomatic Omicron illness after BNT162b2 or ChAdOx1 vaccination [10], recent results on the vaccine effectiveness of Ad26.COV.2 booster dose in South Africa against Omicron hospitalization also demonstrates the value of booster vaccinations [2].

## Data Availability

All data produced in the present study are available upon reasonable request to the authors

## Acknowledgements

The authors would like to express special appreciation to the study participants and all the healthcare workers at Chris Hani Baragwanath Academic Hospital, Charlotte Maxeke Johannesburg Academic Hospital, and Helen Joseph Hospital and to the Wits VIDA staff.

Wits VIDA HCW Study Group: Yasmin Adam, Ziyaad Dangor, David P. Moore, Charl Verwey, Sithembiso C. Velaphi, Firdose Nakwa, Rudo Mathivha, Kavita Makan, Colin Menezes, Merika Tsitsi, Temnewo Habte, Jeremy Nel, William Malebati, Mervin Mer, Adam Mohamed, Rajen Morar, Palesa Motshabi Chakane, Thendo Mpapele, Feroza Motara, Dalubuhle Ndiweni, Debbie White, Farzanah Laher, Shakeel McKenzie, Sihle Mtshali.

## Contributions

MCN: conceptualize the study and the analysis, overall supervision of the project, data interpretation, wrote the first draft of the manuscript. SS: sample analysis, data analysis, manuscript revision; VB: sample analysis, data analysis, manuscript revision; GK: data interpretation, manuscript revision; RA: data analysis, data interpretation, writing. SAM: conceptualize the study, manuscript revision.

## Ethical considerations

The study was approved by the Human Research Ethics Committee of the University of the Witwatersrand (reference number 200405) and conducted in accordance with Good Clinical Practice guidelines. All study participants provided written informed consent.

## References

1. https://www.nicd.ac.za/diseases-a-z-index/disease-index-covid-19/sars-cov-2-genomic-surveillance-update/.

2. Gray, G.E., et al., Vaccine effectiveness against hospital admission in South African health care workers who received a homologous booster of Ad26.COV2 during an Omicron COVID19 wave: Preliminary Results of the Sisonke 2 Study. 2021: p. 2021.12.28.21268436.

3. Nunes, M.C., et al., Severe Acute Respiratory Syndrome Coronavirus 2 Infection Among Healthcare Workers in South Africa: A Longitudinal Cohort Study. Clin Infect Dis, 2021. 73(10): p. 1896–1900.

4. Madhi, S.A., et al., South African Population Immunity and Severe Covid-19 with Omicron Variant. 2021: p. 2021.12.20.21268096.

5. Kwatra, G., et al., Correlation of dried blood spots and plasma for quantification of Immunoglobulin (IgG) against Receptor binding domain and full length spike protein of SARS-CoV-2. J Virol Methods, 2022. 300: p. 114394.

6. Hothorn, T., K. Hornik, and A. Zeileis, Unbiased Recursive Partitioning: A Conditional Inference Framework. Journal of Computational and Graphical Statistics, 2006. 15(3): p. 651–674.

7. Altarawneh, H., et al., Protection afforded by prior infection against SARS-CoV-2 reinfection with the Omicron variant. 2022: p. 2022.01.05.22268782.

8. Dejnirattisai, W., et al., Omicron-B.1.1.529 leads to widespread escape from neutralizing antibody responses. bioRxiv, 2021.

9. Bartsch, Y., et al., Preserved Omicron Spike specific antibody binding and Fc-recognition across COVID-19 vaccine platforms. medRxiv, 2021.

10. Andrews, N., et al., Effectiveness of COVID-19 vaccines against the Omicron (B.1.1.529) variant of concern. 2021: p. 2021.12.14.21267615.

